# Psychiatric comorbid disorders of cognition: A machine learning approach using 1,175 UK Biobank participants

**DOI:** 10.1101/19006189

**Authors:** Chenlu Li, Delia A. Gheorghe, John E.J. Gallacher, Sarah Bauermeister

## Abstract

**Background:** Conceptualising comorbidity is complex and the term is used variously. Here, it is the coexistence of two or more diagnoses which might be defined as ‘chronic’ and, although they may be pathologically related, they may also act independently^1^. Of interest here is the comorbidity of common psychiatric disorders and impaired cognition.

**Objectives:** To examine whether anxiety and/or depression are important longitudinal predictors of cognitive change.

**Methods:** UK Biobank participants used at three time points (n= 502,664): baseline, 1^st^ follow-up (n= 20,257) and 1^st^ imaging study (n=40,199). Participants with no missing data were 1,175 participants aged 40 to 70 years, 41% female. Machine learning (ML) was applied and the main outcome measure of reaction time intraindividual variability (cognition) was used.

**Findings:** Using the area under the Receiver Operating Characteristic (ROC) curve, the anxiety model achieves the best performance with an Area Under the Curve (AUC) of 0.68, followed by the depression model with an AUC of 0.63. The cardiovascular and diabetes model, and the covariates model have weaker performance in predicting cognition, with an AUC of 0.60 and 0.56, respectively.

**Conclusions:** Outcomes suggest psychiatric disorders are more important comorbidities of long-term cognitive change than diabetes and cardiovascular disease, and demographic factors. Findings suggest that psychiatric disorders (anxiety and depression) may have a deleterious effect on long-term cognition and should be considered as an important comorbid disorder of cognitive decline.

**Clinical implications:** Important predictive effects of poor mental health on longitudinal cognitive decline should be considered in secondary and also primary care.

**Summary Box:** *What is already known about this subject? 3-4 bullet points:*

- Poor mental health is associated with cognitive deficits.
- One in four older adults experience a decline in affective state with increasing age.
- ML approaches have certain advantages in identifying patterns of information useful for the prediction of an outcome. What are the new findings? 3-4 bullet points

- Psychiatric disorders are important comorbid disorders of long-term cognitive change.
- Machine-learning methods such as sequence learning based methods are able to offer non-parametric joint modelling, allow for multiplicity of factors and provide prediction models that are more robust and accurate for longitudinal data
- The outcome of the RNN analysis found that anxiety and depression were stronger predictors of change IIV over time than either cardiovascular disease and diabetes or the covariate variables. How might it impact on clinical practice in the foreseeable future?
The important predictive effect of mental health on longitudinal cognition should be noted and, its comorbidity relationship with other conditions such as cardiovascular disease likewise to be considered in primary care and other clinical settings

## Background

Conceptualising comorbidity is complex and the term is used variously. Here, we refer to it as the coexistence of two or more diagnoses which might be defined as ‘chronic’ and, although they may be pathologically related to each other, they may also act independently^1^Of interest here is the comorbidity of common psychiatric disorders and impaired cognition.

Psychiatric disorders such as anxiety and depression (poor mental health) are associated with cognitive deficits^2^, with higher baseline levels of depressive symptoms predicting a steeper decline in delayed recall and global cognition at follow-up^3^, and longitudinal slowing in processing speed^4^. Furthermore, at least one in four older adults experience a gradual decline in affective state^5^ with increasing age, suggesting that along with age-related cognitive decline, there could be a comorbid relationship of unclear temporality^6^. The challenge is to measure this relationship in the most effective way. A cognitive measure which is associated with both longitudinal cognitive decline and poor mental health is reaction time intraindividual variability (RT IIV)^2 7^. RT IIV can be defined as trial-to-trial within-person variability in reaction time across individual trials within a single task for an individual. It is indicative of neurobiological disturbance, an increase of which is associated older age, mild cognitive impairment, dementia and mortality^8^.

Machine learning (ML) approaches have certain advantages in identifying patterns of information useful for the prediction of an outcome, even when complex high-dimensional interactions exist^9^. Unlike hypothesis-driven models, ML approaches are refrained from assumptions on data distributions, especially one of normally distributed independence of attributes or linear data behaviour which are often failed to be met by large registries incorporating biological data. In addition, ML approaches have shown certain advantages in examining potential predictors simultaneously in an unbiased manner. Although ML methods have been used successfully to develop risk-prediction schemes in other areas of medicine, applications to psychiatric comorbid disorders have so far relied on small samples and thin predictor sets, failing to realize the full potential of the methods. With this in consideration, this study applies an ML technique on UK Biobank data to longitudinally examine whether anxiety/depression are important predictors of cognitive change.

The UK Biobank is a large population-based prospective cohort study of 502,664 participants. Invitations to participate in the UK Biobank study were sent to 9.2 million community-dwelling persons in the UK who were registered with the UK National Health Service (NHS) aged between 37 and 73 years. A response rate of 5.5% was recorded. Assessments took place at 22 centres across the UK where participants completed undertook comprehensive mental health, cognitive, lifestyle, biomedical and physical assessments. Mental health and cognitive assessments were completed on a touchscreen computer^10^. Here we use a ML approach to investigate whether the presence of anxiety and depressive symptoms in UK Biobank leads to greater cognitive change measured from baseline to follow-up.

## Objective

To examine whether anxiety and/or depression are important longitudinal predictors of cognitive change.

## Methods

### Governance

Ethical approval was granted to Biobank from the Research Ethics Committee - REC reference 11/NW/0382^10^. All analyses were conducted using data from UK Biobank application 15008.

### Population sample

The study considered for inclusion, the baseline sample of 502,664 UK Biobank participants aged between 37 and 73 years. For the analysis, participants with mental health and cognitive data at three time periods (recruitment, 5 year follow-up, and the 1st imaging sub-study) were used. For the purposes of this analysis the imaging sub-study data represents approximately a 10 year follow-up period from recruitment. Participants who chose to respond, ‘I don’t know’ or ‘I do not wish to answer’ to items within either of the scales (depression and neuroticism) items at the three time periods were excluded, respectively (leaving 373,210 participants at baseline, 129,468 at first assessment and 3,501 at imaging sub-study). Casewise deletion was applied to all demographic variables (2,326 were excluded). All analyses were conducted listwise and the number of participants with no missing data across all measurement variables of interest were n = 1,175.

### Assessment

Details of the UK Biobank assessment procedures may be found elsewhere^10^. Cognitive performance was assessed using a ‘stop-go’ task where RT IIV was used as a sensitive measure of cognitive change with greater RT IIV over time indicating decline in performance^7^. Mental health was assessed using two items from the Patient Health Questionnaire nine-item scale (PHQ-9)^11^ to measure depression (PHQ-2: ‘Over the last 2 weeks, how often have you been bothered by little interest or pleasure in doing things?’; ‘Over the last 2 weeks, how often have you been bothered by feeling down, depressed, or hopeless?’)^12^ and the 12-item Eysenck Personality Questionnaire-Revised (EPQ-R) Neuroticism scale as a proxy measure of anxiety^13 14^ The other measures relevant to this analysis included self-reported cardiovascular disease (CVD: including heart attack, angina and stroke) and diabetes, and observed body mass index (BMI) as indicators of wider co-morbidity^15 16^ Self-reported smoking, alcohol consumption, and fruit and vegetable consumption were used as indicators of lifestyle. Self-reported education, employment, ethnicity, and household income were used as socioeconomic indicators.

### Analytic strategy

Data were modelled as follows. Both age and RT IIV were modelled as continuous variables. The RT IIV was calculated as the standard deviation of each participant’s RTs over the test trials^17^ Participants with only one valid score at baseline or follow-up were omitted. The IIVs showed log– normal distributions and were natural log transformed. Depressed mood items were modelled as categorical variables (Not at all as 1, Several days as 2, More than half the days as 3 and Nearly every day as 4), and anxiety items were modelled as binary variables (yes/no). CVD and diabetes were modelled as binary indicators (present/absent). Gender was modelled as binary indicator (male/female). BMI was treated as a 3 level factor (normal (<25), overweight (25-29.9), obese (>29.9)). Lifestyle data were largely considered as binary indicators (smoker/non-smoker, <5 portions of fruit and vegetable per day / 5 or more portions per day), however alcohol was treated as a continuous variable (g/day). Socioeconomic data were considered as binary indicators (employed/not employed, college degree/no degree, white ethnicity/other) excepting income which was modelled as a 5 level factor (<£18,000, £18,000 -£30,999, £31,000 - £51,999; £52,000 -£100,000, >£100,000).

### Machine learning model

A recurrent neural network (RNN) is a class of artificial neural networks that is often utilized as an effective prediction tool for longitudinal biomedical data. By exhibiting temporal dynamic behaviour for time sequences, it allows for temporal dependencies between measurements. As a sequence learning based method, it is able to offer non-parametric joint modelling of longitudinal data^18 19^. Long short-term memory (LSTM) is an RNN architecture developed to deal with the exploding and vanishing gradient problem during backpropagation through time. It is able to effectively capture long-term temporal dependencies by employing a constant error carousel (memory cell) which maintains its state over arbitrary time intervals, and three nonlinear gating units (input, forget and output) which regulate the information flow into and out of the cell.^18 20-23^. This architecture is applied to longitudinally model the prediction abilities of psychiatric comorbidity disorders on cognition via sequence-to-sequence learning.

The dataset was partitioned into two non-overlapping subsets: 75% of the within-class subjects for training and 25% for testing. Data were rescaled [0 and 1] to meet the default hyperbolic tangent activation function of the LSTM model. The Adam version of stochastic gradient descent algorithm was applied to tune the weights of the network. This optimisation algorithm combines the advantages of Adaptive Gradient Algorithm (AdaGrad) and Root Mean Square Propagation (RMSProp), thus being effective in handling sparse gradients on non-stationary problems. Furthermore, it compares favourably to other stochastic optimization methods in practice^24^. Hyperparameters are optimised on the training set, e.g., the model was set to fit 1000 training epochs with a batch size set to the number of available training subjects. Mean absolute error (MAE) and Area under the Receiver Operating Characteristic (ROC) Curve (AUC) were used to assess model performance, respectively. Permutation tests were applied to obtain *p*-values of AUC. To evaluate whether depression and anxiety are important comorbid disorders of cognition and, to compare the prediction abilities of diabetes and cardiovascular disease the model was run systematically to include the following as features:

I. Covariates only as reference model (age; BMI; lifestyle factors; gender; socioeconomic factors)
II. Neuroticism scale + reference model
III. Depression scale + reference model
IV. Diabetes and cardiovascular disease + reference model

Exposure (outcome) was cognition (RT IIV). Given that depression mood disorder and anxiety have previously been considered to be comorbid disorders of impaired cognition, we expected predictions based on comorbid mental health disorders to outperform predictions based only on covariate variables.

A complete data analysis approach was used and all analyses were conducted on the Dementias Platform UK Data Portal^25^ in Python 3.6.7 and MATLAB R2019.

## Findings

Complete data were available for 1,175 Participants (Table 1). The sample was aged between 40 and 70 years at baseline, 41% female, and relatively healthy with few participants showing comorbidities and few reporting unhealthy behaviour such as smoking (5%).

**Table 1.**
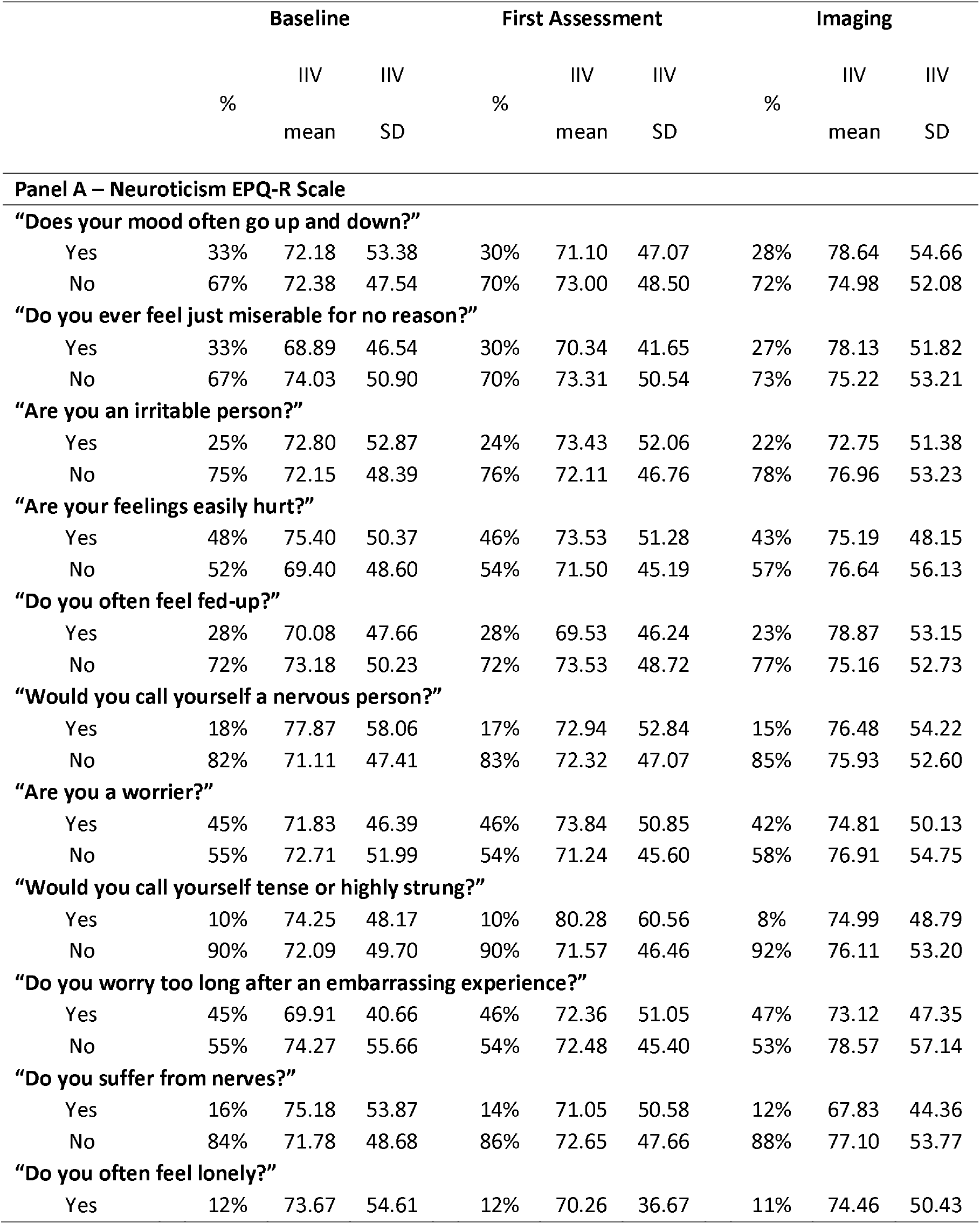
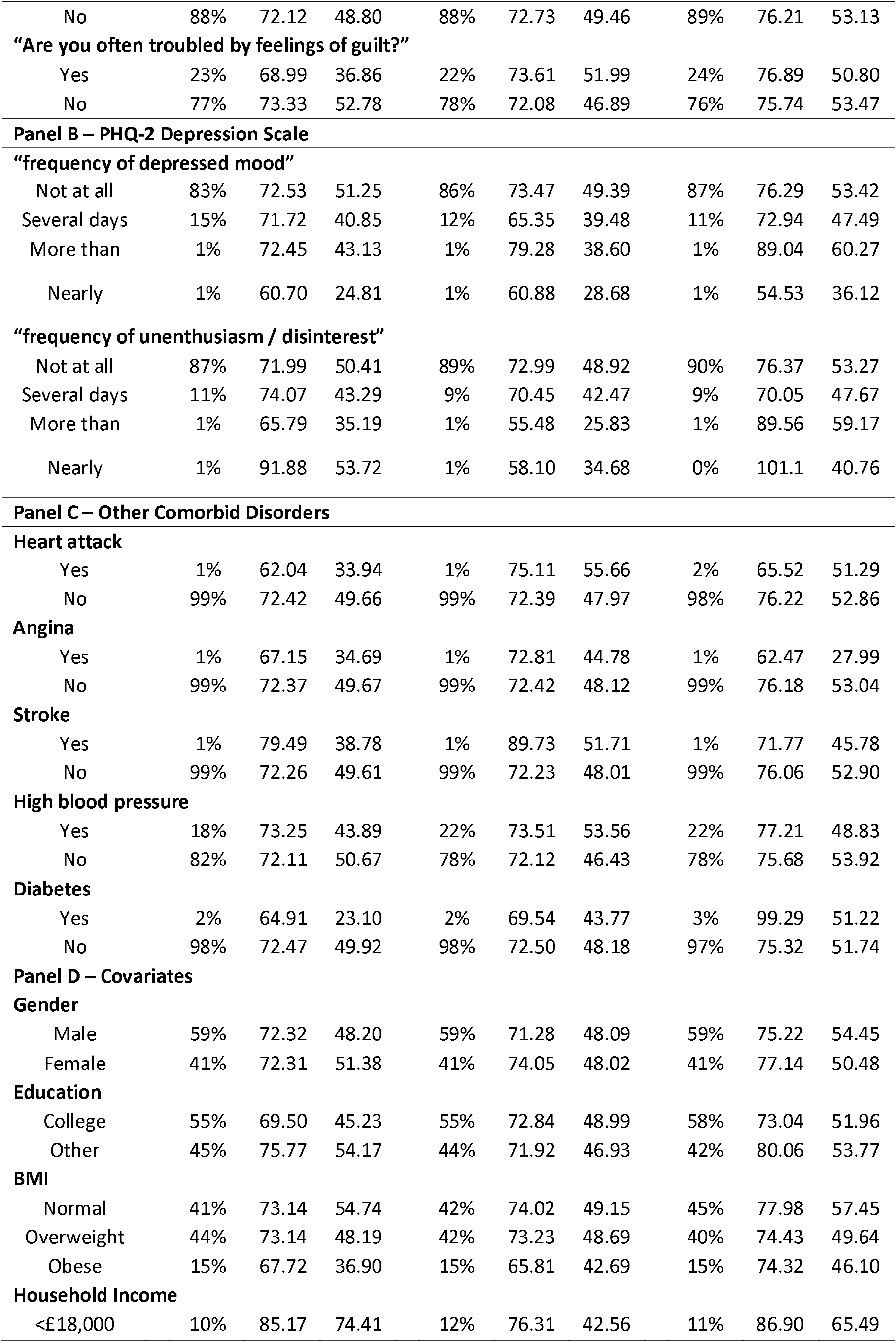
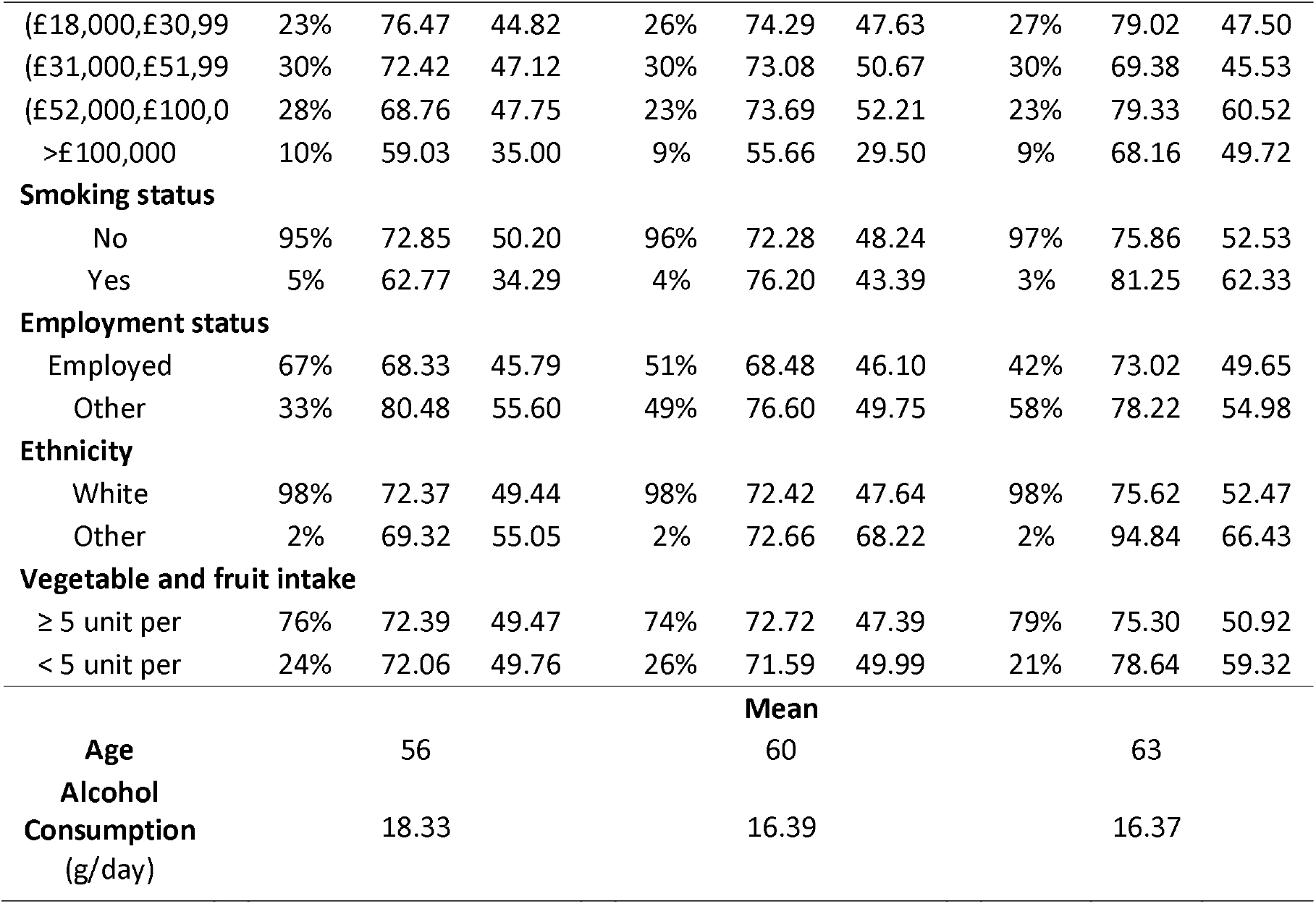
Descriptive statistics.

|  | Baseline |  |  | First Assessment |  |  | Imaging |  |  |
| --- | --- | --- | --- | --- | --- | --- | --- | --- | --- |
|  | % | IIV<br>mean | IIV<br>SD | % | IIV<br>mean | IIV<br>SD | % | IIV<br>mean | IIV<br>SD |
| Panel A – Neuroticism EPQ-R Scale |  |  |  |  |  |  |  |  |  |
| “Does your mood often go up and down?” |  |  |  |  |  |  |  |  |  |
| Yes | 33% | 72.18 | 53.38 | 30% | 71.10 | 47.07 | 28% | 78.64 | 54.66 |
| No | 67% | 72.38 | 47.54 | 70% | 73.00 | 48.50 | 72% | 74.98 | 52.08 |
| “Do you ever feel just miserable for no reason?” |  |  |  |  |  |  |  |  |  |
| Yes | 33% | 68.89 | 46.54 | 30% | 70.34 | 41.65 | 27% | 78.13 | 51.82 |
| No | 67% | 74.03 | 50.90 | 70% | 73.31 | 50.54 | 73% | 75.22 | 53.21 |
| “Are you an irritable person?” |  |  |  |  |  |  |  |  |  |
| Yes | 25% | 72.80 | 52.87 | 24% | 73.43 | 52.06 | 22% | 72.75 | 51.38 |
| No | 75% | 72.15 | 48.39 | 76% | 72.11 | 46.76 | 78% | 76.96 | 53.23 |
| “Are your feelings easily hurt?” |  |  |  |  |  |  |  |  |  |
| Yes | 48% | 75.40 | 50.37 | 46% | 73.53 | 51.28 | 43% | 75.19 | 48.15 |
| No | 52% | 69.40 | 48.60 | 54% | 71.50 | 45.19 | 57% | 76.64 | 56.13 |
| “Do you often feel fed-up?” |  |  |  |  |  |  |  |  |  |
| Yes | 28% | 70.08 | 47.66 | 28% | 69.53 | 46.24 | 23% | 78.87 | 53.15 |
| No | 72% | 73.18 | 50.23 | 72% | 73.53 | 48.72 | 77% | 75.16 | 52.73 |
| “Would you call yourself a nervous person?” |  |  |  |  |  |  |  |  |  |
| Yes | 18% | 77.87 | 58.06 | 17% | 72.94 | 52.84 | 15% | 76.48 | 54.22 |
| No | 82% | 71.11 | 47.41 | 83% | 72.32 | 47.07 | 85% | 75.93 | 52.60 |
| “Are you a worrier?” |  |  |  |  |  |  |  |  |  |
| Yes | 45% | 71.83 | 46.39 | 46% | 73.84 | 50.85 | 42% | 74.81 | 50.13 |
| No | 55% | 72.71 | 51.99 | 54% | 71.24 | 45.60 | 58% | 76.91 | 54.75 |
| “Would you call yourself tense or highly strung?” |  |  |  |  |  |  |  |  |  |
| Yes | 10% | 74.25 | 48.17 | 10% | 80.28 | 60.56 | 8% | 74.99 | 48.79 |
| No | 90% | 72.09 | 49.70 | 90% | 71.57 | 46.46 | 92% | 76.11 | 53.20 |
| “Do you worry too long after an embarrassing experience?” |  |  |  |  |  |  |  |  |  |
| Yes | 45% | 69.91 | 40.66 | 46% | 72.36 | 51.05 | 47% | 73.12 | 47.35 |
| No | 55% | 74.27 | 55.66 | 54% | 72.48 | 45.40 | 53% | 78.57 | 57.14 |
| “Do you suffer from nerves?” |  |  |  |  |  |  |  |  |  |
| Yes | 16% | 75.18 | 53.87 | 14% | 71.05 | 50.58 | 12% | 67.83 | 44.36 |
| No | 84% | 71.78 | 48.68 | 86% | 72.65 | 47.66 | 88% | 77.10 | 53.77 |
| “Do you often feel lonely?” |  |  |  |  |  |  |  |  |  |
| Yes | 12% | 73.67 | 54.61 | 12% | 70.26 | 36.67 | 11% | 74.46 | 50.43 |
| No | 88% | 72.12 | 48.80 | 88% | 72.73 | 49.46 | 89% | 76.21 | 53.13 |
| **“Are you often troubled by feelings of guilt?”** |  |  |  |  |  |  |  |  |  |
| Yes | 23% | 68.99 | 36.86 | 22% | 73.61 | 51.99 | 24% | 76.89 | 50.80 |
| No | 77% | 73.33 | 52.78 | 78% | 72.08 | 46.89 | 76% | 75.74 | 53.47 |
| **Panel B – PHQ-2 Depression Scale** |  |  |  |  |  |  |  |  |  |
| **“frequency of depressed mood”** |  |  |  |  |  |  |  |  |  |
| Not at all | 83% | 72.53 | 51.25 | 86% | 73.47 | 49.39 | 87% | 76.29 | 53.42 |
| Several days | 15% | 71.72 | 40.85 | 12% | 65.35 | 39.48 | 11% | 72.94 | 47.49 |
| More than | 1% | 72.45 | 43.13 | 1% | 79.28 | 38.60 | 1% | 89.04 | 60.27 |
| Nearly | 1% | 60.70 | 24.81 | 1% | 60.88 | 28.68 | 1% | 54.53 | 36.12 |
| **“frequency of unenthusiasm / disinterest”** |  |  |  |  |  |  |  |  |  |
| Not at all | 87% | 71.99 | 50.41 | 89% | 72.99 | 48.92 | 90% | 76.37 | 53.27 |
| Several days | 11% | 74.07 | 43.29 | 9% | 70.45 | 42.47 | 9% | 70.05 | 47.67 |
| More than | 1% | 65.79 | 35.19 | 1% | 55.48 | 25.83 | 1% | 89.56 | 59.17 |
| Nearly | 1% | 91.88 | 53.72 | 1% | 58.10 | 34.68 | 0% | 101.1 | 40.76 |
| **Panel C – Other Comorbid Disorders** |  |  |  |  |  |  |  |  |  |
| **Heart attack** |  |  |  |  |  |  |  |  |  |
| Yes | 1% | 62.04 | 33.94 | 1% | 75.11 | 55.66 | 2% | 65.52 | 51.29 |
| No | 99% | 72.42 | 49.66 | 99% | 72.39 | 47.97 | 98% | 76.22 | 52.86 |
| **Angina** |  |  |  |  |  |  |  |  |  |
| Yes | 1% | 67.15 | 34.69 | 1% | 72.81 | 44.78 | 1% | 62.47 | 27.99 |
| No | 99% | 72.37 | 49.67 | 99% | 72.42 | 48.12 | 99% | 76.18 | 53.04 |
| **Stroke** |  |  |  |  |  |  |  |  |  |
| Yes | 1% | 79.49 | 38.78 | 1% | 89.73 | 51.71 | 1% | 71.77 | 45.78 |
| No | 99% | 72.26 | 49.61 | 99% | 72.23 | 48.01 | 99% | 76.06 | 52.90 |
| **High blood pressure** |  |  |  |  |  |  |  |  |  |
| Yes | 18% | 73.25 | 43.89 | 22% | 73.51 | 53.56 | 22% | 77.21 | 48.83 |
| No | 82% | 72.11 | 50.67 | 78% | 72.12 | 46.43 | 78% | 75.68 | 53.92 |
| **Diabetes** |  |  |  |  |  |  |  |  |  |
| Yes | 2% | 64.91 | 23.10 | 2% | 69.54 | 43.77 | 3% | 99.29 | 51.22 |
| No | 98% | 72.47 | 49.92 | 98% | 72.50 | 48.18 | 97% | 75.32 | 51.74 |
| **Panel D – Covariates** |  |  |  |  |  |  |  |  |  |
| **Gender** |  |  |  |  |  |  |  |  |  |
| Male | 59% | 72.32 | 48.20 | 59% | 71.28 | 48.09 | 59% | 75.22 | 54.45 |
| Female | 41% | 72.31 | 51.38 | 41% | 74.05 | 48.02 | 41% | 77.14 | 50.48 |
| **Education** |  |  |  |  |  |  |  |  |  |
| College | 55% | 69.50 | 45.23 | 55% | 72.84 | 48.99 | 58% | 73.04 | 51.96 |
| Other | 45% | 75.77 | 54.17 | 44% | 71.92 | 46.93 | 42% | 80.06 | 53.77 |
| **BMI** |  |  |  |  |  |  |  |  |  |
| Normal | 41% | 73.14 | 54.74 | 42% | 74.02 | 49.15 | 45% | 77.98 | 57.45 |
| Overweight | 44% | 73.14 | 48.19 | 42% | 73.23 | 48.69 | 40% | 74.43 | 49.64 |
| Obese | 15% | 67.72 | 36.90 | 15% | 65.81 | 42.69 | 15% | 74.32 | 46.10 |
| **Household Income** |  |  |  |  |  |  |  |  |  |
| <£18,000 | 10% | 85.17 | 74.41 | 12% | 76.31 | 42.56 | 11% | 86.90 | 65.49 |
| (£18,000,£30,99 | 23% | 76.47 | 44.82 | 26% | 74.29 | 47.63 | 27% | 79.02 | 47.50 |
| (£31,000,£51,99 | 30% | 72.42 | 47.12 | 30% | 73.08 | 50.67 | 30% | 69.38 | 45.53 |
| (£52,000,£100,0 | 28% | 68.76 | 47.75 | 23% | 73.69 | 52.21 | 23% | 79.33 | 60.52 |
| >£100,000 | 10% | 59.03 | 35.00 | 9% | 55.66 | 29.50 | 9% | 68.16 | 49.72 |
| **Smoking status** |  |  |  |  |  |  |  |  |  |
| No | 95% | 72.85 | 50.20 | 96% | 72.28 | 48.24 | 97% | 75.86 | 52.53 |
| Yes | 5% | 62.77 | 34.29 | 4% | 76.20 | 43.39 | 3% | 81.25 | 62.33 |
| **Employment status** |  |  |  |  |  |  |  |  |  |
| Employed | 67% | 68.33 | 45.79 | 51% | 68.48 | 46.10 | 42% | 73.02 | 49.65 |
| Other | 33% | 80.48 | 55.60 | 49% | 76.60 | 49.75 | 58% | 78.22 | 54.98 |
| **Ethnicity** |  |  |  |  |  |  |  |  |  |
| White | 98% | 72.37 | 49.44 | 98% | 72.42 | 47.64 | 98% | 75.62 | 52.47 |
| Other | 2% | 69.32 | 55.05 | 2% | 72.66 | 68.22 | 2% | 94.84 | 66.43 |
| **Vegetable and fruit intake** |  |  |  |  |  |  |  |  |  |
| ≥ 5 unit per | 76% | 72.39 | 49.47 | 74% | 72.72 | 47.39 | 79% | 75.30 | 50.92 |
| < 5 unit per | 24% | 72.06 | 49.76 | 26% | 71.59 | 49.99 | 21% | 78.64 | 59.32 |
| **Mean** |  |  |  |  |  |  |  |  |  |
| **Age** |  | 56 |  |  | 60 |  |  | 63 |  |
| **Alcohol Consumption (g/day)** |  | 18.33 |  |  | 16.39 |  |  | 16.37 |  |

The outcome of the RNN analysis found that anxiety and depression were stronger predictors of change IIV over time than either cardiovascular disease and diabetes or the covariate variables, as measured by the ROC curve (Figure 1 and Table 2). The AUC curve was 0.68 (*p* < 0.0001) for anxiety, followed by the depression model with an AUC of 0.63 (*p* < 0.0001). The cardiovascular and diabetes model and the demographics model had weaker performance with an AUC of 0.60 (*p* = 0.0036) and 0.56 (*p* = 0.1126), respectively. The anxiety model prospectively identified 63.24% of patients who eventually reached high IIV (i.e., sensitivity), and 53.35% of patients who maintained low IIV (i.e., specificity). Correspondingly, the anxiety model had a positive predictive value (PPV) of 52.72%, and a negative predictive value (NPV) of 63.68%. The depression model achieved slightly smaller yet still significant (*p* < 0.0001) predictive performance in comparison with the other models. The RNNs results suggest that the anxiety and depression model outperformed the reference model (Model I – covariates only). In general, the neuroticism and depression scales helped improve the accuracy in capturing the pattern of IIV, and therefore may be more important predictors of cognitive change over cardiovascular disease and diabetes. The results also suggest increased risk of multiple mental health disorders occurring together, suggesting depression and anxiety are potential comorbid disorders of cognition.

**Table 2.**
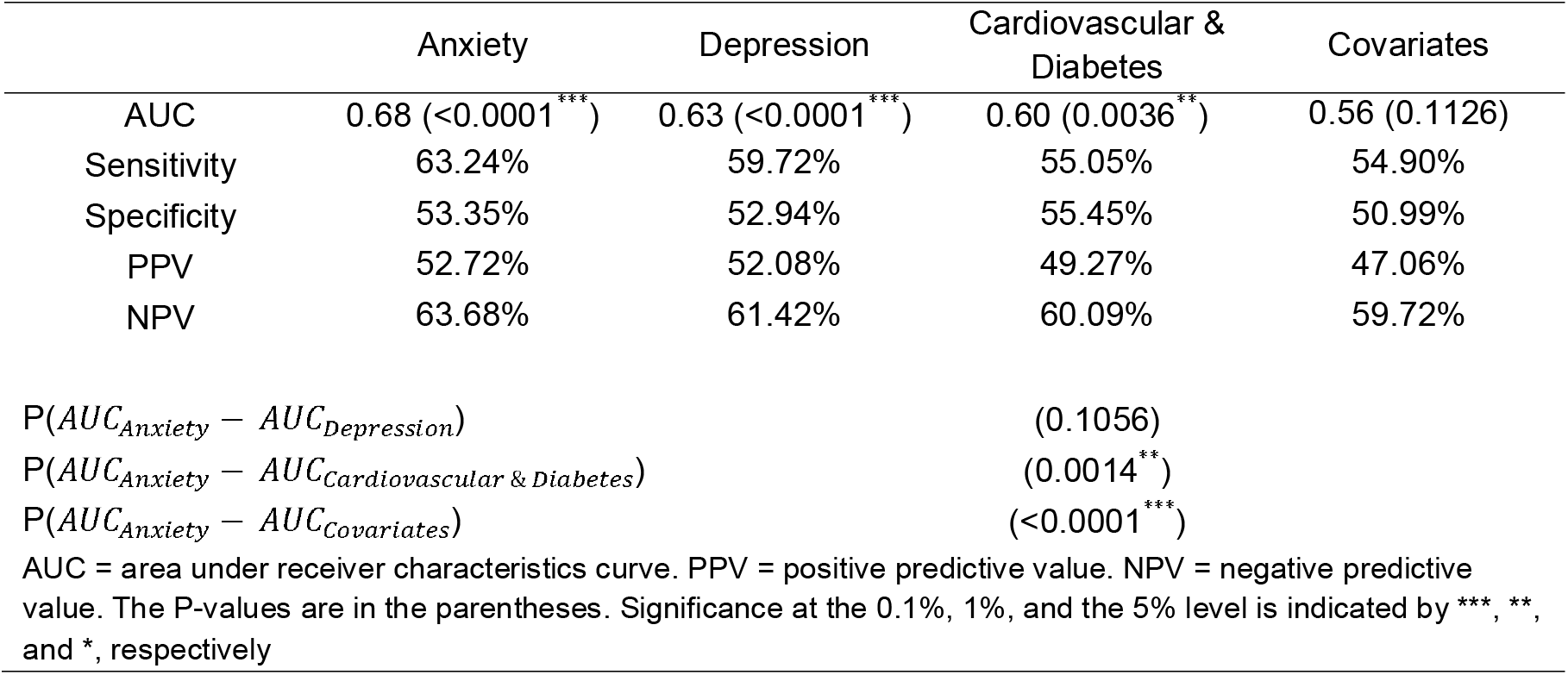
Model performance for comorbid longitudinal predictors of IIV.

**Figure.**
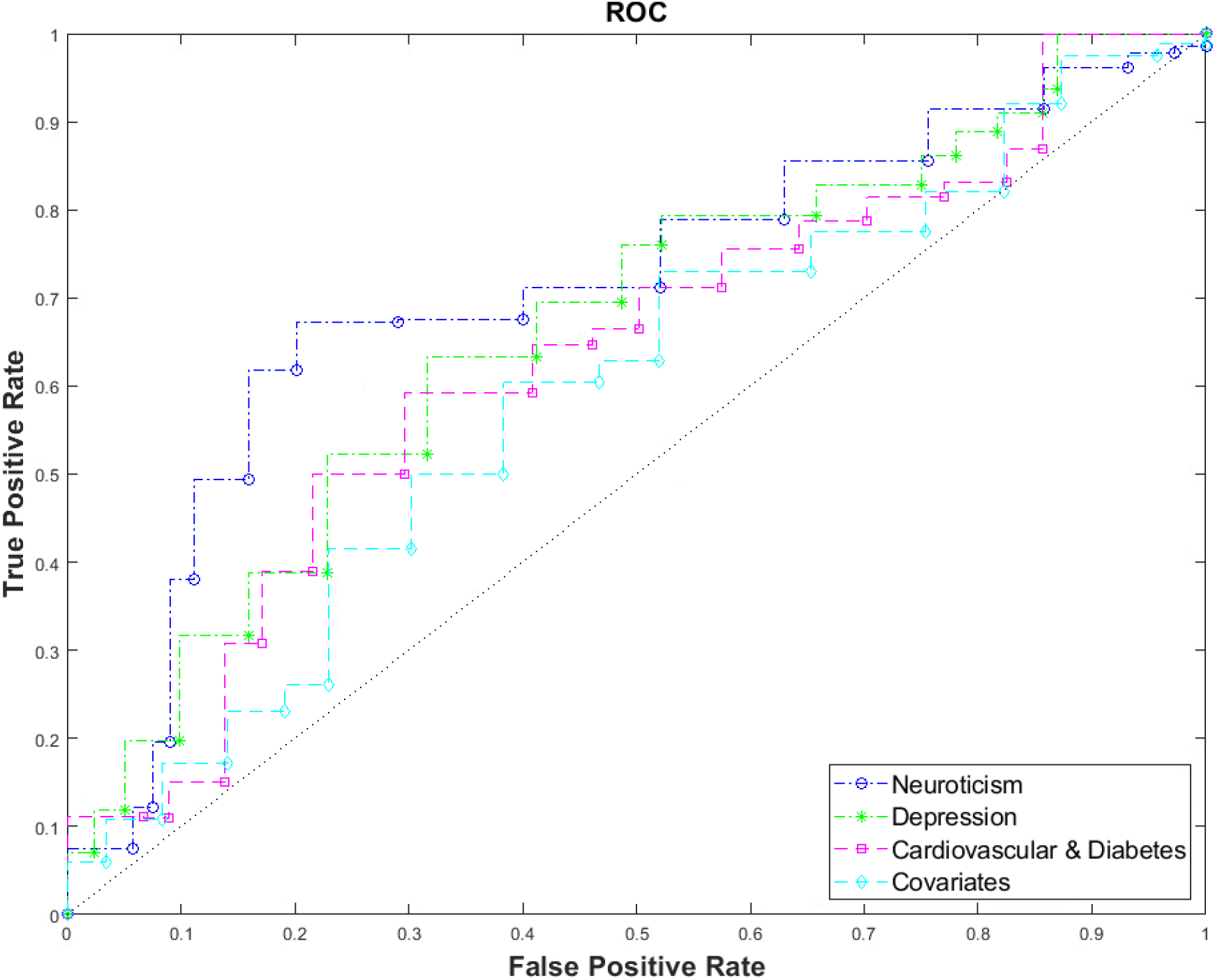
ROCs for RNN Models I to IV.

## Discussion

A longitudinal analysis of the comorbid predictors of mental health disorders on cognitive change was conducted using ML in a relatively healthy population sample. Evidence was found suggesting that anxiety and depression are important comorbid disorders of cognition but when cardiovascular disease, diabetes and other covariate predictors were included in the model these were not as important on long-term effect. This suggests that poor mental health has a significant deleterious effect on long-term cognition, and may be considered an important comorbid disorder of cognitive decline. The contribution of the present investigation is thereby three-fold.

First, this study used the community-based sample from UK Biobank with a general population large enough for identification of valid effects which might be implied across clinical and research settings. Clinical-based samples have been criticised for inconsistency in diagnoses when patients are treated over time in a variety of facilities or at different times in the same facility, or for failure to report all the comorbid diagnoses of a patient^26^. In comparison, community-based samples are based on a system in which a practical number of criteria are rated, or collected and assessed on every patient, providing unbiased prevalence or incidence rates of comorbidity, or unbiased estimates of risk factors for comorbidity^26^.

Second, this study investigates the psychiatric comorbid disorders of cognition in a comprehensive ML design which was applied to examine if psychiatric disorders as measured by self-report scales are important predictors of impaired cognition. Unlike hypothesis-driven statistical techniques, the sequence learning based method is able to offer non-parametric joint modelling, allow for multiplicity of factors and provide prediction models that are more robust and accurate for longitudinal data^18^.

Third, this study is conducted longitudinally. Disease progression modelling using longitudinal data may provide clinicians with improved tools for diagnosis and monitoring of diseases. We apply the widely used LSTM network to capture the long-term temporal dependencies among measurements without making parametric assumptions about cognitive trajectories. To our knowledge there are no studies which have employed the ML techniques presented here to investigate the comorbidity of mental health disorders and longitudinal cognitive decline using cardiovascular disease and diabetes as comorbidity covariates. Utilising ML methodologies for clinical data such as psychiatric outcomes may provide additional information which is not possible with traditional statistical methodologies. In future research, approaches used here may include imputation on missing data in other longitudinal cohorts. For instance, by pre-processing data interpolation using mean or forward imputation, or utilizing possible correlations between missing values’ patterns and the target^27^.

There are general limitations to our work which should be noted. Participants in UK Biobank are selected from a general healthy population and outcomes may not reflect implications for those with clinically diagnosed psychiatric disorders of anxiety and depression. Here, also, measures of psychological state at point of assessment have been used to measure poor mental health (anxiety and depression). A further limitation is that only one cognitive measure has been used but findings would benefit from application across measures of wider cognition, e.g., memory and executive function.

### Clinical Implications

The implication of this work is that the important predictive effect of mental health on longitudinal cognition should be noted,^28^ and its comorbid relationship with other conditions such as cardiovascular disease likewise to be considered in primary care and other clinical settings.

## Data Availability

All data is available on the Dementias Platform UK Data Portal after permission from UK Biobank

https://portal.dementiasplatform.uk/

## Acknowledgements

This is a DPUK supported project with all analyses conducted on the DPUK Data Portal, constituting part 1 of DPUK Application 0132.

The Medical Research Council supports DPUK through grant MR/L023784/2. CL was funded by DPUK to complete this analysis.

## Authors’ Contributions

CL, JG and SB conceptualised the idea. CL analysed the data, CL, DG, JG and SB interpreted the data, CL wrote the 1^st^ draft of the manuscript. DG and SB commented, edited and proofread the manuscript. All authors read and approved the final manuscript.

